# Neural consequences of 5-Hz transcranial alternating current stimulation over right hemisphere: an eLORETA EEG study

**DOI:** 10.1101/2023.10.09.23296743

**Authors:** Tien-Wen Lee, Chiang-Shan R. Li, Gerald Tramontano

## Abstract

**Introduction:** Transcranial alternating current stimulation (tACS) at 5-Hz to the right hemisphere can effectively alleviate symptoms of anxiety. The objective of this study was to explore the neural mechanisms that drive the therapeutic benefits.

**Methods:** We collected electroencephalography (EEG) data from 24 participants with anxiety disorders both before and after the tACS treatment during a single session. We applied the stimulation over the right hemisphere, with 1.0 mA at F4, 1.0 mA at P4, and 2.0 mA at T8, following the 10-10 EEG convention. With eLORETA, we transformed the scalp signals into the current source density in the cortex. We then assessed the differences between post- and pre-treatment brain maps across multiple spectra (delta to low gamma) with non-parametric statistics.

**Results:** We observed a trend of heightened power in alpha and reduced power in mid-to-high beta and low gamma, in accord with the EEG markers of anxiolytic effects reported in previous studies. Additionally, contrary to the widely circulated entrainment theory of the neural effects of tACS, we observed a consistent trend of de-synchronization at the stimulating sites across spectra.

**Conclusion:** We confirmed that tACS 5-Hz over the right hemisphere demonstrated EEG markers of anxiety reduction. Regarding changes in power spectra, the effects of tACS on the brain are intricate and cannot be explained solely by entrainment theory.

## Introduction

Transcranial alternating current stimulation (tACS) influences cortical excitability and activity (Antal et al., 2008; Bland and Sale, 2019). With the resurgence of transcranial electrical stimulation (tES) as a tool for basic and clinical research (Guleyupoglu et al., 2013), studies have also focused on tACS as a treatment option for neuropsychiatric conditions (Alexander et al., 2019; Clancy et al., 2017). Neural oscillation is one of the hallmarks of brain activities (Lee, 2016). By implementing alternating currents, investigators may employ tACS to manipulate neural rhythmicity and synchronize oscillations within the brain (Ali et al., 2013).

tACS has been proposed as a treatment option for anxiety disorders (Clancy et al., 2017; Lee and Tramontano, 2023). Applying alpha frequency tACS at 2.0 mA to the occipito-parietal sites, Clancy et al. reported a rapid reduction in anxious arousal and aversion to auditory stimuli (Clancy et al., 2017). Lee et al. reported that 5-Hz tACS at 2.0 mA over the right hemisphere with the currents oscillating between T8 and F4/P4 (10-10 EEG convention) alleviated anxiety symptom severity (Lee and Tramontano, 2023). However, the neural mechanisms underlying the therapeutic efficacy of tACS remain unclear.

Compared with transcranial direct current stimulation (tDCS), which has also been used to treat anxiety disorders (Hampstead et al., 2016), tACS does not directly affect neuronal membrane potential or its propensity to hyperpolarize or depolarize (Nitsche and Paulus, 2000). Supported by combined EEG and tACS research probing alpha and low gamma frequencies (Helfrich et al., 2014; Voss et al., 2014), the theory of entrainment suggests that when a specific frequency of tACS is applied, neural oscillations in the targeted brain region(s) are synchronized to match the stimulation frequency.

The current study aimed to test this hypothesis. We recorded EEGs before and after one tACS treatment session in the experiment design. In contrast to prior tACS studies that targeted a single area with alternating current, we used a montage that covered frontal, parietal, and temporal areas (Alexander et al., 2019; Clancy et al., 2017). This design enabled the assessment of the effects of tACS on multiple brain regions. Further, unlike previous studies of electrode-based analysis (Alexander et al., 2019; Clancy et al., 2017), we converted the scalp-recorded EEG signals by exact Low-Resolution Electromagnetic Tomography (eLORETA)—from topography to tomography. Our brain-based, as opposed to electrode-based, approach would unveil the global effects of 5-Hz tACS on the brain.

Studies have found that adults who experience stress, anxiety, and excessive rumination may present higher levels of fast beta waves in the EEG (Díaz et al., 2019; Engel and Fries, 2010; Spitzer and Haegens, 2017), and the situations that cause worry and social anxiety can increase gamma power (Miskovic et al., 2010; Oathes et al., 2008). In contrast, relaxation and restfulness have long been associated with higher alpha power (Berger, 1929; Sharma and Singh, 2015). Thus, given the calming effects of 5-Hz tACS, we anticipate that the treatment will cause a rise in alpha power and a decline in beta and gamma powers in the brain (*Prediction One*). Furthermore, as per the entrainment theory of tACS, theta power is expected to undergo broad enhancement (*Prediction Two*).

## Materials and Methods

### Participants

This study focused on patients with anxiety disorders who received 5-Hz tACS treatment over the right hemisphere. We reviewed the data collected from our clinics between 2018 and 2022 after obtaining approval from a private Review Board (Pearl IRB; https://www.pearlirb.com/). EEG changes following tES were the main foci of this research. Thirty-two anxiety patients who had pre- and post-treatment EEG were identified. The anxiety reduction was around 60%. As for detailed treatment efficacy and clinical profiles, please refer to another report (Lee and Tramontano, 2023).

### Administration of tACS

We used Starstim-8, an FDA-approved neuromodulation device developed by Neuroelectrics, Inc., to treat anxiety disorders. The electrodes, connected via wires to a rechargeable battery, transmit electric currents directly to the scalp and brain. Before the electrodes were attached, the scalp was scrubbed with skin preparation gel. The conductive gel was then applied to fill the space between the scalp and electrodes. These procedures ensured optimal contact of the electrodes with the scalp and kept the impedance level < 5 k ohms (DaSilva et al., 2011).

The montage of electrodes covered the right lateral side of the head at F4, T8, and P4 positions in terms of the 10-10 EEG convention. The peak current intensity for T4 was 2.0 mA, while those at F4 and P4 were 1.0 mA. Alternating sinewave currents oscillated at 5 Hz between electrodes T4 and electrodes F4 and P4. During the neuromodulation session, the subjects were asked to practice progressive muscle relaxation by watching a standardized video to enhance the treatment effects (Jacobson, 1938). The setup of tACS is illustrated in Figure 1.

**Fig. 1.** An illustration of the neuromodulation setup. Three electrode positions were wire-connected to a rechargeable battery: F4, P4, and T8.

To help the patients to become accustomed to the tES, a dose escalation strategy was adopted and was completed in the first three sessions: 1.0 mA for session 1, 1.5 mA for session 2, and 2.0 mA for session 3. The stimulation lasted 25 minutes, with a 1-minute ramp-up and a 30-second ramp-down phase to reduce skin irritation.

### EEG recording and data pre-processing

The treatment dosage of tES generally was stabilized at session 3 or 4, in case the patient required more exposure to get accustomed to the dosage. Before and after the 3^rd^ (or 4^th^) neuromodulation session, the Brainmaster device (https://brainmaster.com/) was used to acquire 10 min eye-open digital EEG data at 256 samples/sec with linked-ear reference.

We employed the software EEGLAB to edit the EEG traces (Delorme and Makeig, 2004). Data preprocessing included band-pass filtering (1–50 Hz), automatic artifact removal (Artifact Subspace Reconstruction), and manual elimination of the remaining noisy portions. A single rater (TW Lee) carried out the above processing. With the artifacts, including those associated with blinks and eye movements, removed, the cleaned EEG data were segmented into 2-sec epochs and imported to eLORETA for subsequent analyses.

### eLORETA and statistical analyses

A tomographic method for the analyses of neural electric activity, eLORETA, improved over standardized LORETA by incorporating optimized leadfield weights, enabled precise localization of deeper structures (Jurcak et al., 2007; Pascual-Marqui, 2007). In contrast to the parametric approach, the eLORETA linear imaging method provides a weighted minimum norm inverse solution, where the weights are adaptive to the data (i.e., data-dependent) to accommodate measurement and biological noises. Any arbitrary point-test sources can be correctly computed with exact, zero-error localization. Based on the principles of linearity and superposition, eLORETA is suitable for delineating distributed electric sources (current source density; CSD) in the brain cortex, albeit with low spatial resolution. The power spectrum of delta (1–4 Hz), theta (4–8 Hz), alpha (8–12 Hz), mid-to-high beta (15–30 Hz), and low gamma (30–45 Hz) were derived (Lee and Tramontano, 2022). With eLORETA, the neural informatics from the electrodes can be projected to a Talairach brain template with 6,239 gray matter voxels.

The CSDs were calculated for each voxel and each frequency band. We adopted two normalization strategies, i.e., subject- and frequency-wise. Assign the CSD value for a voxel at a frequency band as s(b,v), where b and v respectively denote a frequency band and a voxel. Subject-wise normalization scales s(v,b) with a grand average over all frequencies and voxels, 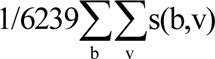, with 6239 total cortical voxels. The denominator of frequency-wise normalization is 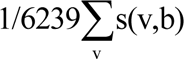 for each frequency band. Subject-wise normalization is standard in neuroimaging analysis intending to discount variation due to non-experimental session effect (e.g., in the routine analysis of functional magnetic resonance imaging data (Cox, 1996)), while frequency-wise normalization procedure may provide additional, complementary information. For example, if the voxels of an area showed increased CSD but some voxels within that area showed decreased CSD after frequency-wise normalization, the finding indicates that the enhancement of CSD is lower in that sub-region. In other words, the enhancement is not evenly distributed across the area.

The comparisons of post- and pre-treatment cortical CSDs were performed by paired t-tests and a non-parametric statistical method (SnPM) with 5,000 times randomization (Nichols and Holmes, 2002; Westfall and Young, 1993). The max-statistics and corrected *P*-values for multiple testing can thus be derived. The exceedance proportion test was used to evaluate the significance of activity based on its spatial extent, revealing clusters of supra-threshold voxels. These statistical modules and normalization schemes were implemented in the eLORETA software. The calculation based on subject-wise normalized CSD data was the primary analysis. The null hypothesis was that the CSDs showed no differences after tES treatment at a *P* value of 0.05, two-tailed.

## Results

Twenty-four of the 32 patients (75%) had pre- and post-tACS EEGs that met quality assessment. The 24 patients ranged from 15.2 to 54.0 with a mean ± SD = 34.6 ± 14.2 years in age and comprised 9 males and 15 females. All 24 tolerated the maximum tES current at 2.0 mA.

### Statistics based on subject-wise normalization (primary analysis)

The neural responses to 5-Hz tACS exhibited apparent spectral effects, with a notable global decrease in CSD in the delta and gamma ranges and a concurrent increase in CSD in the alpha spectrum. Mixed patterns were observed in the theta and mid-to-high beta CSDs, with attenuation in the right hemisphere. The *P* values of the peak statistics after multiple corrections were not significant but approached significance for the more lenient 1-tailed comparisons at delta, alpha, and gamma frequencies. Exceedance proportion tests disclosed that when the brain map was thresholded at 2.66 and 3.10, the *P* value of spatial extent (cluster distribution) was 0.037 and 0.016, respectively. Except for the comparison at the theta band, all the other frequency maps survived the statistical tests of spatial extents (two-tailed). The results are illustrated in Figure 2, and the statistics of regional peaks are summarized in Table 1 (left column).

**Fig. 2.**
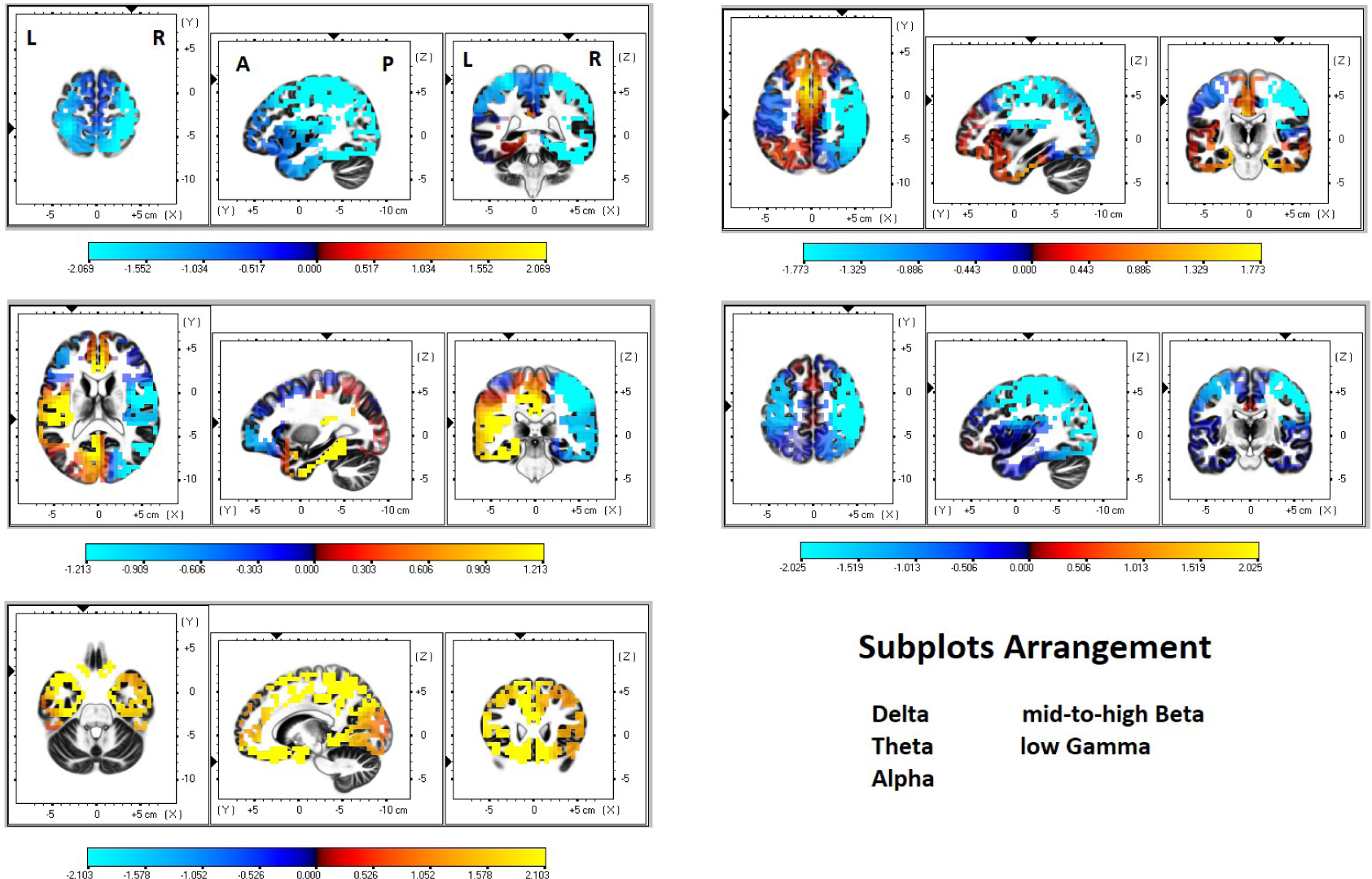
The changes in CSD under subject-wise normalization (post-treatment minus pre-treatment) are displayed through the five defined spectra (delta, theta, alpha, mid-to-high beta, and low gamma), with warm colors indicating an increase and cool colors indicating a decrease. The axial (left), sagittal (middle), and coronal (right) sections are centered around the highest statistics. The color bar is included at the end of each sub-graph and is scaled at 50% maximum to illustrate better the trend of CSD changes for each frequency band.

**Table 1.**
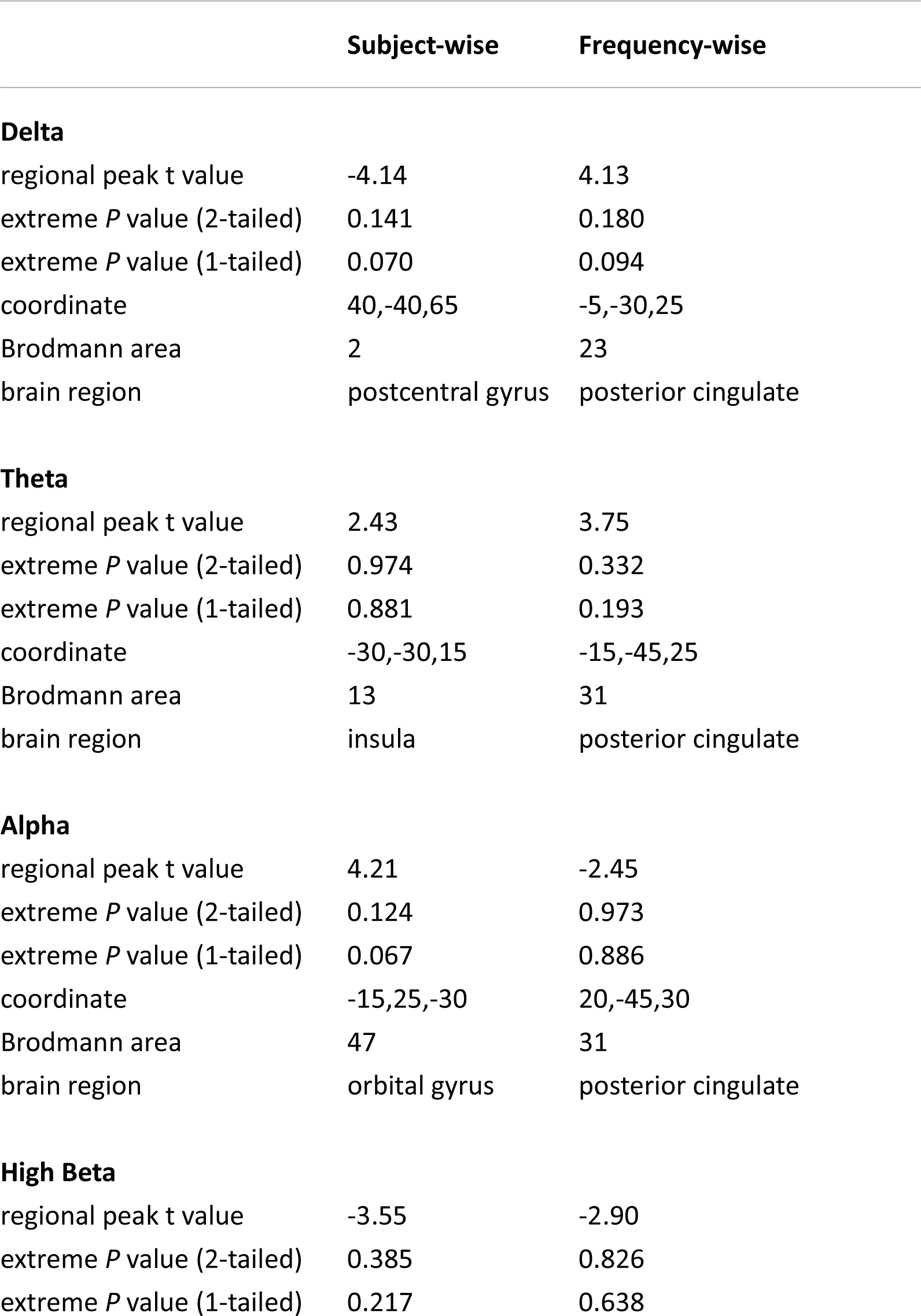
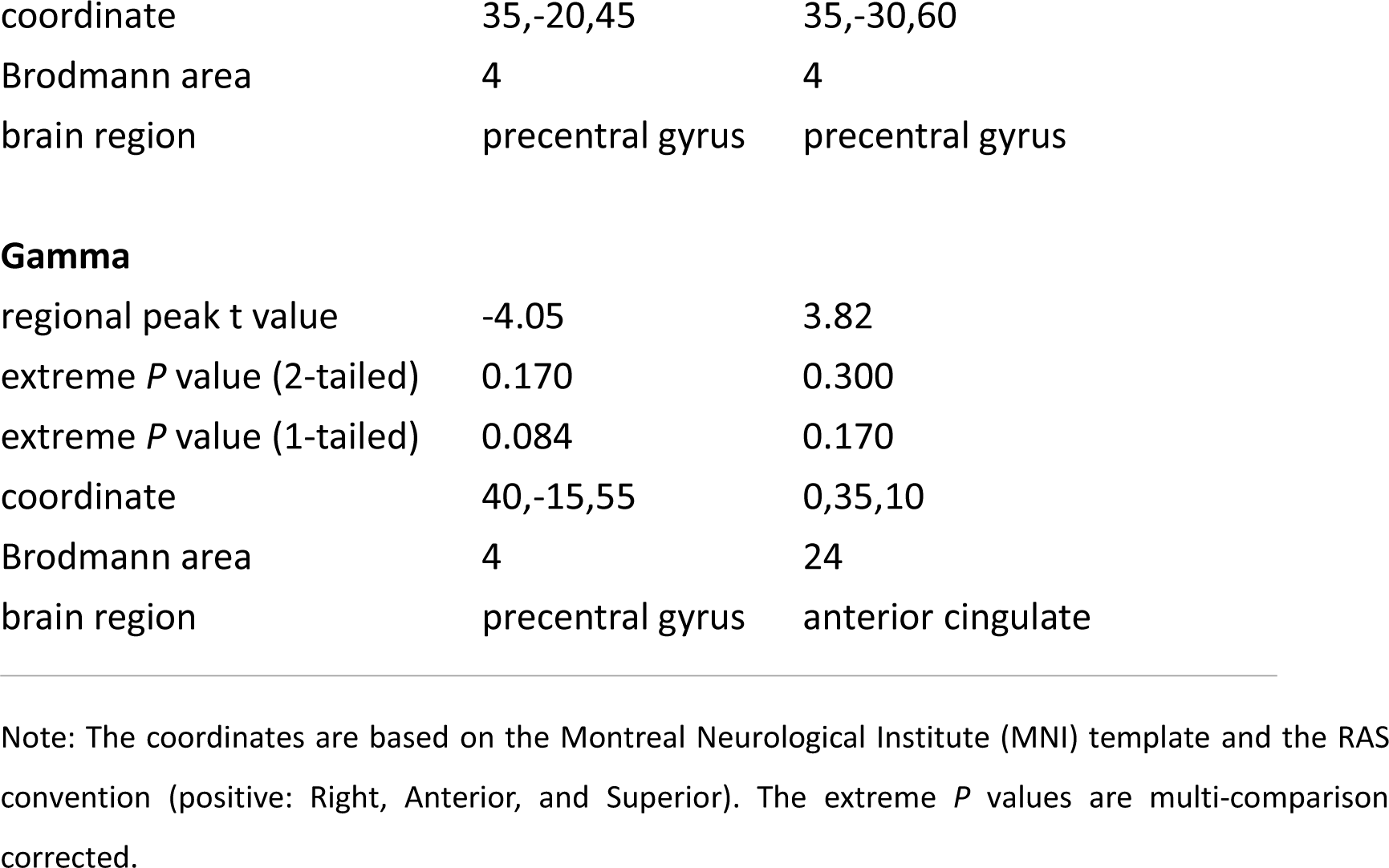
Summary of neural changes to neuromodulation treatment based on subject-wise and frequency-wise normalizations, with the points with highest t values reported.

### Statistics based on frequency-wise normalization (supplementary analysis)

After frequency-wise normalization, the differential brain maps showed similar trends across spectra. Generally, the changes in the right hemisphere were negative except in the temporal lobe, frontal pole, and inferior frontal cortex) and, in contrast, those over the midline/vertex of the brain were positive. As for the left hemisphere, the directionality of changes was relatively negative for the mid-to-high beta and low gamma maps. Mixed patterns were noticed for delta, theta, and alpha counterparts. Figure 3 shows these findings. As for the lateral temporal cortex, corresponding to position T8, negative values were noticed for delta and theta, and positive values for the remaining spectrum. The results centering around the points with max statistics are illustrated in Figure 4 and summarized in Table 1 (right column; not significant).

**Fig. 3.**
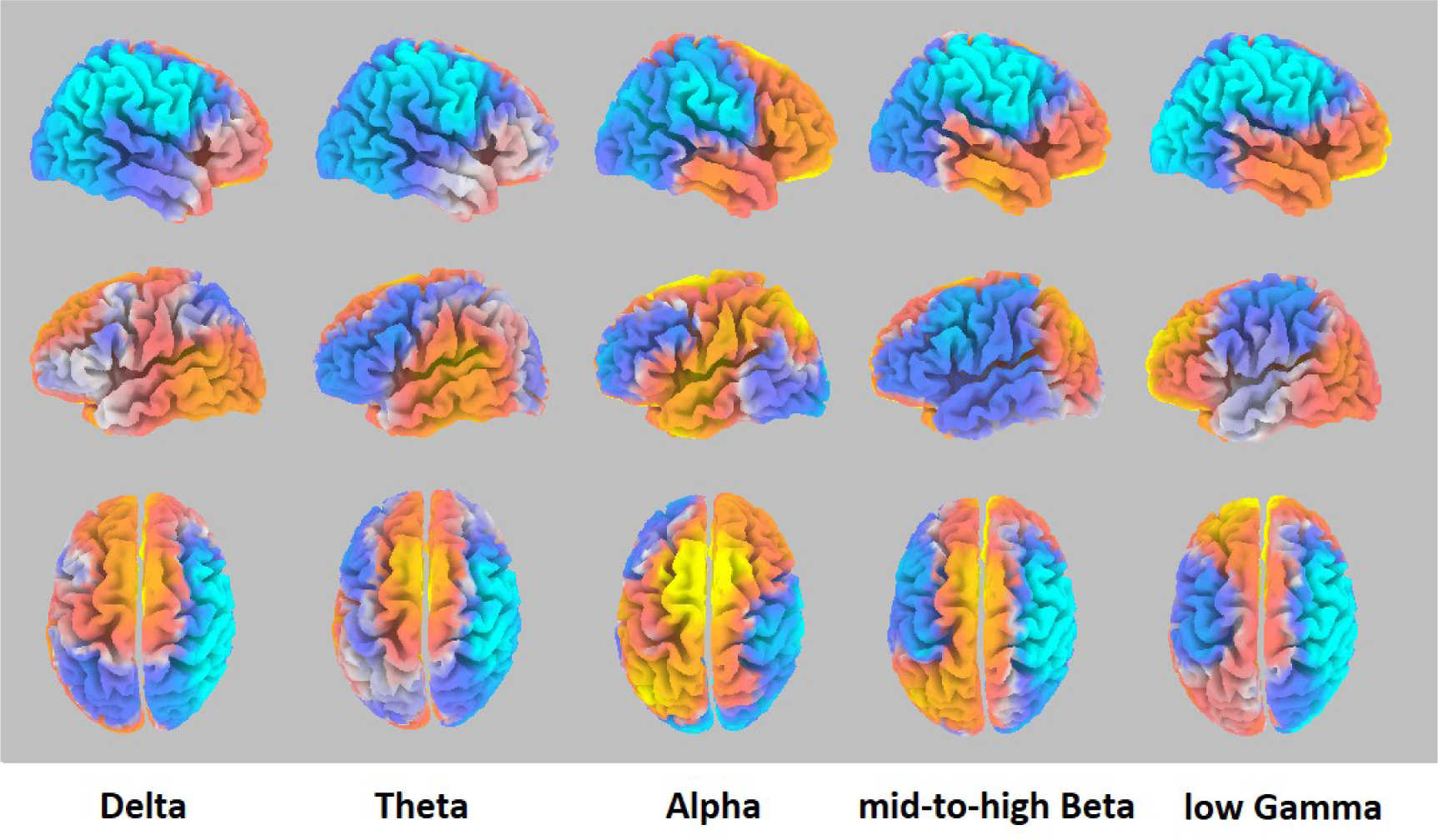
Lateral and top views of the CSD differences under frequency-wise normalization (post-treatment minus pre-treatment), highlighting the trends of changes across different spectra. Upper row: right hemisphere. Middle row: left hemisphere. Lower row: top view. The color bars can be referred to in Figure 4.

**Fig. 4.**
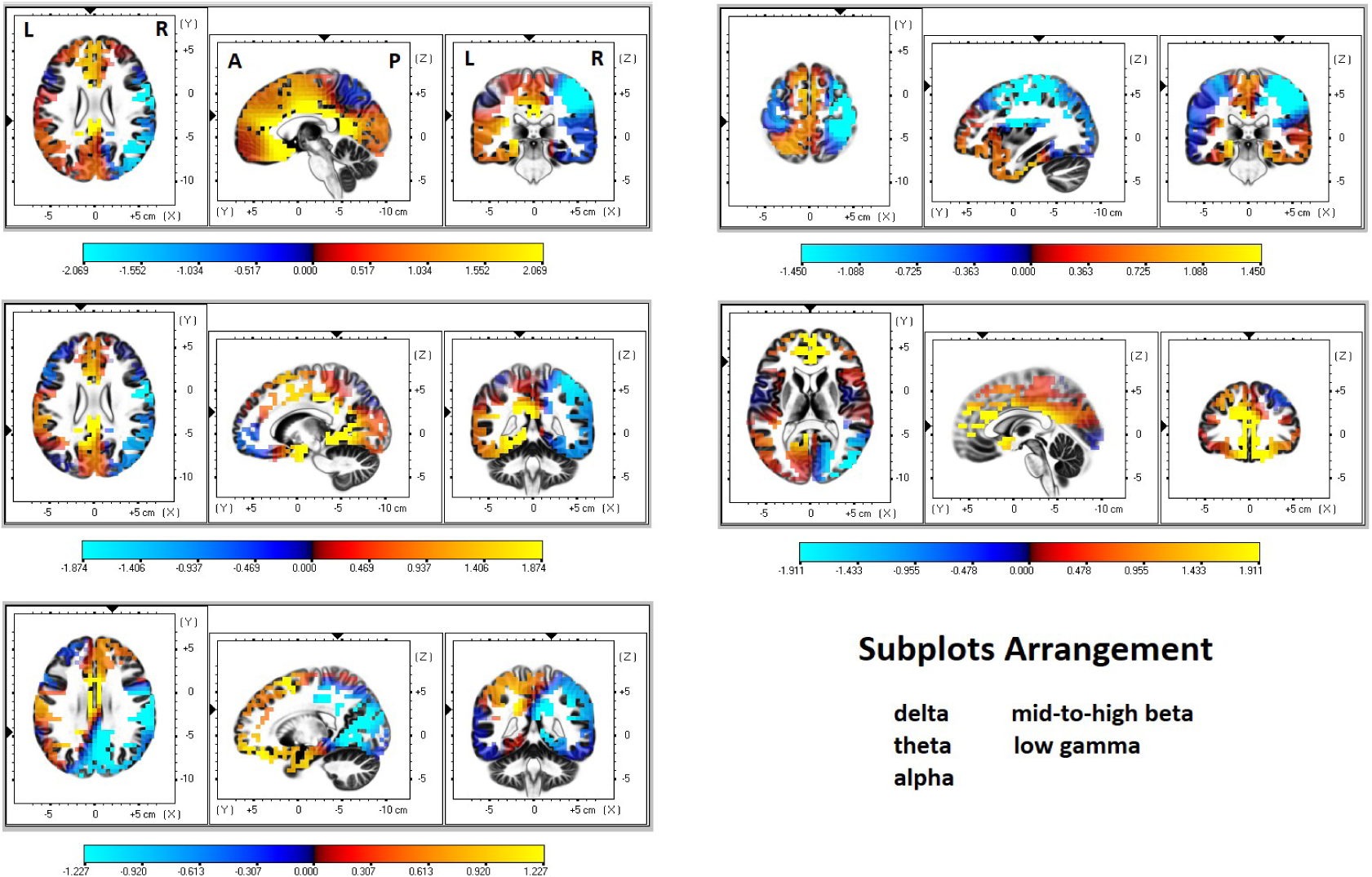
The changes in CSD under frequency-wise normalization (post-treatment minus pre-treatment) are displayed through the five selected spectra (delta, theta, alpha, mid-to-high beta, and low gamma), with warm colors indicating an increase and cool colors indicating a decrease. The axial (left), sagittal (middle), and coronal (right) sections are centered around the highest statistics. The color bar is included at the end of each sub-graph and is scaled at 50% maximum to illustrate better the trend of CSD changes for each frequency band.

The spatial extents did not surpass the exceedance proportion tests.

## Discussion

We studied the neural consequences of tACS at 5-Hz and 2.0 mA over the right scalp, with a tripod design aimed at easing anxiety (Lee and Tramontano, 2023). The montage covered frontal, parietal, and temporal regions; thus, the effects of tACS can be viewed as a hemispheric-level modulation. Pre- and post-treatment EEGs were collected and analyzed for 24 participants using an imaging method eLORETA to solve the inverse EEG problem of source localization. SnPM framework was adopted to provide statistical inferences. The regional peak values failed to surpass or, at most, approached significance, but the spatial extent of thresholded clusters (exceedance proportion tests) survived *P* < 0.05, rejecting the null hypothesis. The results were consistent with the impression of tES that its immediate influence was mild and widespread (Antal et al., 2008), with the latter well demonstrated by simulation studies (Manoli et al., 2012; Saturnino et al., 2017). In accord, it is more appropriate to interpret the findings in terms of large-scale (i.e., lobar, system, or hemispheric) rather than focal based on peak statistics or Brodmann area, as in the convention of source localization of event-related potentials. In addition, 5-Hz tACS not only modulated the neural responses in the theta range but also triggered robust cross-spectral interactions from delta to gamma bands. In the below, we discussed the main findings from three different perspectives: (1) association with anxiety reduction, which nicely supports *Prediction One*, (2) cross-spectral effects of 5-Hz tACS, which did not agree with *Prediction Two*, and (3) large-scale neurophysiological mechanisms of tACS.

### Neural changes associated with anxiety reduction

Anxiety reduction has been one of the earliest and most common applications of tES, such as Electrosleep and Alpha-Stim (Morriss et al., 2019; Robinovitch, 1914). We invented a novel neuromodulation protocol to treat anxiety disorders, and the symptom severity reduction may reach 60% after 12 treatment sessions (Lee and Tramontano, 2023), consistent with an earlier report (Clancy et al., 2017). The primary eLORETA analyses revealed two findings that align well with the current knowledge of neural markers of anxiety reduction, namely enhancement in alpha and decrease in beta and gamma powers (Berger, 1929; Engel and Fries, 2010; Miskovic et al., 2010; Oathes et al., 2008; Sharma and Singh, 2015; Spitzer and Haegens, 2017). The concordance with the therapeutic effects endorses the validity of the eLORETA results. In the next session, we will integrate and elaborate on the detailed spectral changes obtained through subject-wise and frequency-wise normalizations.

### Spectrum-specific changes to 5-Hz tACS neuromodulation -delta spectrum

There was a trend of global attenuation of delta power (from the subject-wise normalization, we will skip the notes in the following paragraphs). Nevertheless, the reduction was not evenly distributed across the brain, most prominent at the right hemisphere (frontoparietal) where the tACS was applied, then the left parietal region, least at the vertex and limbic cortex, including midline structure (supplemented by results from frequency-wise normalization, we will skip the notes in the following paragraphs).

### -theta spectrum

Neural changes at the theta range were only partly similar to those of delta. The power decrease in the right hemisphere (frontoparietal) was the consistent finding. On the contrary, the left hemisphere and the midline structures showed a trend of increased power. Our *Prediction Two,* inferred from entrainment theory, was not supported. Together with the findings at delta frequency, our results provide several insights into the physiological reactions of tACS. First, in contrast to entrainment theory, 5-Hz tACS interferes with or desynchronizes the targeted cortical rhythm. Second, the interference is more noticeable for the spectrum at and lower than the stimulating frequency (i.e., delta, lower than 5-Hz) rather than in the higher range (i.e., alpha, higher than 5-Hz; detailed below). Third, the contralateral enhancement in power may work through the mechanism of interhemispheric rivalry, a common observation in the field of tES research (Hilgetag et al., 2001; Kinsbourne, 1974; Naeser et al., 2005). We want to address a caveat here that the above-summarized patterns may or may not repeat themselves for the tACS at other frequencies, which requires further studies to examine.

### -alpha spectrum

There was a global trend of enhanced Alpha power after 5-Hz tACS treatment, which sharply contrasts the picture of delta. The magnitude of change was not evenly distributed across different brain regions. The increase was most prominent in the left hemisphere and midline region and less in the stimulating side (right hemisphere) and visual cortex. As described above, the increased alpha power is believed to be related to anxiolytic effects. The finding cannot be explained by entrainment theory, either. Given that the energy expenditure of the brain is relatively stable (Lee, 2016), the enhanced alpha could be the consequence of the attenuated delta, also named “net zero-sum” by some authors (Brem et al., 2014).

### -mid-to-high beta and low gamma spectra

Since the power change maps of the mid-to-high beta and low gamma were similar, we discussed them together. The difference between the two brain maps lay in the midline and vertex regions, where the mid-to-high Beta map showed increased power, see Figure 2. We noticed a trend of decreased spectral power of the frequency from 15 to 45 Hz, which was linked to anxiety reduction, as discussed above. Again, the power decrease was more remarkable in the right hemisphere, the same side as the tACS application. The neurophysiological underpinning needs to be clarified. We speculated that the observed power decrease at higher frequency could be the offset of the increased power at alpha and beta 1 (12 to 15 Hz, not reported). Nevertheless, the results provided preliminary evidence that the neural modulatory effect by 5-Hz tACS can go beyond its default frequency and reach the gamma range.

### Large-scale neural responses to tACS

Previous work suggested that tACS may influence neuronal firing rate and timing (Anastassiou et al., 2010; Krause et al., 2019; Radman et al., 2009; Radman et al., 2007). Further, tACS could affect neural activity through several possible mechanisms, such as stochastic resonance, entrainment of network patterns, and so on (Antal et al., 2017; Antal and Paulus, 2013). From the perspective of large-scale network functions, our analyses revealed novel insights into the effects of tACS on brain dynamics, as elaborated below.

First, our attempt to apply entrainment theory to large-scale networks and the spectra pertinent to physiology generally failed. When the neural effects were suppressive, as shown in the primary analysis with subject-wise normalization, there was a more substantial reduction in activity in the right hemisphere, specifically in the delta, theta, mid-to-high beta, and low gamma frequencies. Conversely, when the effects were elevating, the increases in activity in the right hemisphere were less noticeable, particularly in the alpha frequency. We thus concluded that introducing artificially generated narrow-band current to the brain was unfavorable to the underlying neural oscillations, with its influences going beyond the default frequency. Previous research already challenged the entrainment theory in its account of large-scale network findings. For example, Alexander and colleagues employed alpha tACS to target the dorsolateral prefrontal cortex in individuals with major depressive disorder, with the sham condition and 40 Hz stimulation as controls (Alexander et al., 2019). The results showed more responders in the 10 Hz-tACS group, but the alpha power was reduced due to the stimulation. Brignani et al. found that delivering tACS at 6 Hz and 10 Hz over the occipito-parietal area impaired performance in the detection task, echoing that injecting narrow-band artificial current may interfere with rather than facilitate the underneath neural processing (Brignani et al., 2013). Lafton et al. applied the intracranial recording and observed no sleep rhythm entrainment to tACS (Lafon et al., 2017). The decrease in anxiety that occurred alongside the “negative” modulation of the right hemisphere through tACS appeared to echo the emotion lateralization theory (Ross, 2021). More research is necessary to comprehend the connections.

Second, the neural consequences of tACS are far more complicated than expected. Our analyses demonstrated that the changes in brain dynamics involved several well-documented neural interaction mechanisms. The differential CSD map of the theta frequency (primary analysis) and those derived from frequency-wise normalization (supplementary analysis) highlighted the engagement of inter-hemispheric rivalry, as shown in the delta, theta, and alpha maps (Hilgetag et al., 2001; Kinsbourne, 1974; Naeser et al., 2005). The interference from tACS might initiate compensatory cascades of power boost/diminution of the neighboring spectra, possibly mediated by the “net zero-sum” principle to maintain constant energy expenditure (Brem et al., 2014; Lee, 2016). Third, the directionalities of power changes in the midline/limbic structures and the outer cortex were dissociable, in agreement with their distinctive evolutionary and developmental trajectories (Mega et al., 1997). This pattern can readily be appreciated from the results derived from frequency-wise normalization, indicating that the neural reactions to the tACS were system dependent. Further, it was also noted that the changes in CSD in the right temporal region, where T8 tACS was applied, were sometimes consistent (delta, theta, alpha, gamma) and sometimes in the opposite direction (mid-to-high beta) relative to those of the frontoparietal network. The mixed manifestation could reflect that the T8 tACS may modulate the outer temporal cortex, part of it also belongs to the default-mode network, and the inner limbic system at the same time (Lee and Xue, 2018) and hence perturbs the cortico-limbic regulation. These findings highlight the benefits of a brain-based approach in revealing network dynamics that may not otherwise be observed with its electrode-based counterpart.

A concern for the decreases of the delta, beta, and gamma powers is whether tACS at 5-Hz would impair attention and other cognitive functions. Our clinical report addressed this issue and showed no impact of tACS on cognitive capabilities after 12 sessions of treatment (Lee and Tramontano, 2023).

## Limitations and Conclusion

In a protocol to study the effects of tACS on anxiety, we investigated the neural effects of 5-HZ tACS at 2.0 mA over the right hemisphere (F4, P4, T8) by comparing EEG recordings before and after treatment. To substantiate brain-based analyses, the signals captured from the scalp were converted to brain patterns using eLORETA. The resulting neural changes were consistent with what is expected when anxiety is reduced. However, the power analysis results have invalidated the popular entrainment theory. The effects of tACS on the brain are complex and influenced by multiple factors. These include that tACS causes de-synchronization across broad spectra, how different brain networks and hemispheres interact, and the fundamental principles that govern the brain’s functioning. We acknowledged several limitations. First, the electrode number of the 10-20 EEG system is small. However, since both EEG and tACS are low spatial resolution tools, we do not expect much benefit from a higher electrode number. Second, the results present in this study reflect the session effect. Further research is needed to clarify whether comparing brain maps between after a treatment course (say, the 16^th^ treatment session) and baseline (before treatment) yields the same results.

## Authors Contributions

All authors contributed intellectually to this work. TW Lee carried out the analysis and wrote the first draft. All authors revised and approved the final version of the manuscript.

## Data Availability

All data produced in the present study are available upon reasonable request to the authors

## Acknowledgments

This work was supported by NeuroCognitive Institute (NCI) and NCI Clinical Research Foundation Inc. We would like to thank Almeida Sergio for helping prepare the research material.

## Financial support

N/A.

## Statements and Declarations

All authors declare no conflicts of interest.

## Compliance with ethical standards

This research analyzed the databank collected from 2018 to 2022. The authors assert that all procedures contributing to this work comply with the ethical standards of the relevant national and institutional committees on human experimentation and with the Helsinki Declaration of 1975, as revised in 2008.

## Notes

### Competing Interest Statement

The authors have declared no competing interest.

### Funding Statement

No research fund

### Author Declarations

Ethics committee of Pearl IRB gave ethical approval for this work (category: exempt because the study is about analyzing record data)

